# Who Makes It Through the Funnel? Sociodemographic Drivers of Recruitment Completion into an AI-Ready Digital Speech Bank

**DOI:** 10.1101/2025.08.18.25333909

**Authors:** Peyman Nejat, Ashley D. Bachman, Vicki M. Stubbs, Joseph R. Duffy, John L. Stricker, Vitaly Herasevich, David T. Jones, Rene L. Utianski, Hugo Botha

## Abstract

**Background:** Digital recruitment methods offer promising opportunities to address persistent challenges in clinical research participation, particularly in specialized fields like neurology. However, the impact of digital approaches across different socioeconomic and demographic groups remains inadequately understood. This study analyzed participant recruitment pathways in a digital neurology research study to identify sociodemographic factors associated with participation outcomes.

**Methods:** We conducted a longitudinal analysis of 5,846 patients invited to participate in a remote speech capture study for neurological disease research between March and July 2024. Using data from Qualtrics, PTrax, and our recording platform, we tracked participant progression through multiple recruitment checkpoints. Socioeconomic status was assessed using the Housing-based Socioeconomic Status (HOUSES) index and Area Deprivation Index (ADI). We examined associations between participation pathways and demographic factors including age, sex, geographic location, and socioeconomic indices using Kruskal-Wallis and Wilcoxon rank-sum tests.

**Results:** Only 415 participants (7.1%) completed all study requirements. Participants from neighborhoods with higher socioeconomic disadvantage (higher ADI national ranks) were significantly less likely to express interest in initial invitations (median ADI 45.0 vs. 42.0 for responders, *p*<0.001). Urban participants completed enrollment faster than those from rural areas or urban clusters (median 32.0 days vs. 41.0 and 40.0 days, *p*=0.011). Contrary to expectations, younger participants were more likely to drop out at multiple recruitment stages, with the median age increasing from 63 years in the invited cohort to 66.3 years among completers. Female participants required more time to complete enrollment compared to males (median 38.5 days vs. 32.0 days, *p*=0.010). While neighborhood-level socioeconomic status significantly influenced participation, individual housing circumstances showed no significant association across recruitment stages.

**Conclusions:** Digital recruitment methods in neurological research do not automatically overcome traditional barriers to participation and may introduce new disparities related to the digital divide. The significant associations between participation outcomes and sociodemographic factors—particularly neighborhood socioeconomic status, geographic location, age, and sex—highlight the need for targeted recruitment strategies. Researchers should implement multi-channel approaches, design age-specific engagement strategies, address geographic disparities, and consider socioeconomic factors to enhance the inclusivity and effectiveness of digital recruitment in neurological research.

## Introduction

Participant recruitment is a critical component of medical research studies, and the rate of participation varies significantly depending on the type of study, the setting, and the population involved. As an example, recent data suggests that enrollment rate in cancer trials is around 6.3% to 7.1%^1,2^. Enrollment rates may also differ among different populations; specifically, they can be lower among minority, pediatric, and geriatric populations^3,4^. Poor recruitment often results in underpowered studies, which lack the necessary sample size to detect meaningful differences between distinct groups. This can lead to statistically non-significant results even when there are clinically relevant effects^5^.

The rise of digital methods offers a promising avenue to address these recruitment challenges. Digital tools, such as social media platforms, mobile applications, and web-based programs, have expanded the reach of research recruitment efforts. These platforms allow researchers to target specific demographics and geographic areas, making it easier to engage with heterogenous populations that were previously hard to reach through traditional methods^6–8^. For instance, Facebook has been shown to be an effective tool for recruiting participants in health research, particularly among young and hard-to-reach demographics, offering a more representative sample compared to traditional methods^8^.

Despite the benefits, the use of digital tools in recruitment and enrollment presents challenges, including privacy concerns and the need for informed consent. Researchers must navigate these issues carefully to maintain participant trust and ensure ethical standards are met. Moreover, while digital tools are increasingly used, there is a lack of robust evidence supporting their effectiveness compared to traditional recruitment methods, highlighting the need for further research in this area^9,10^. The integration of digital technologies in recruitment processes also introduces potential sampling biases that can inadvertently exclude certain populations. One of the primary concerns is the “digital divide,” which refers to the disparities in access to digital technologies and the internet. This divide is often more pronounced among disadvantaged groups, including racial and ethnic minorities and the elderly, who may have limited access to the internet and lower digital literacy levels^11–13^.For instance, studies have shown that older adults and African American patients are less likely to use digital health portals compared to their younger and White counterparts, highlighting a significant gap in technology utilization^12^. Similarly, individuals from lower socioeconomic status (SES) neighborhoods often have reduced access to the internet and lower health literacy, which can hinder their ability to engage with digital health technologies effectively^13^. While digital tools have the potential to improve the representativeness of trial participants, there is limited evidence supporting their effectiveness in recruiting underrepresented groups^7^. This underscores the need for targeted interventions and strategies to bridge the digital divide and ensure equitable access to digital health resources^14,15^.

Digital recruitment has become especially popular with the increased interest in artificial intelligence in healthcare, which require large and representative datasets to train. For example, the Bridge2AI-Voice program has the explicit goal to create “an ethically sourced flagship dataset to enable future research in artificial intelligence”^16^. These voice recordings are primarily obtained through a digital recruitment pipeline. Our group has similarly endeavored to obtain a large bank of speech recordings focused on neurological disorders, with the goal of subsequently using these data to train AI models, using a primarily digital recruitment approach. Given the importance of understanding the effect digital recruitment has on the sociodemographic distribution of the resulting dataset, our ongoing study offered the ideal setting to formally investigate the pathways participants took during a digital recruitment workflow and where they got lost in the process, examining potential associations with socioeconomic and demographic factors. By identifying and addressing potential factors that may limit the recruitment in similar research, this study aims to ensure equitable access to research studies and the development of interventions that benefit all populations.

## Methods

### Speech Capture Study

As part of an ongoing study to remotely capture speech from patients with neurologic diseases, most neurology patients on our campus are invited to complete a self-administered speech examination. Patient identification is conducted using our Electronic Health Record (Epic). Subsequently, invitations to complete an eligibility survey are sent via the patient portal to interested participants using Qualtrics (https://www.qualtrics.com). The inclusion/exclusion criteria are simple – we invite US based adults (18 and older) who have English listed as a language they can communicate in via spoken language. The eligibility survey additionally explores their understanding of the study through simple yes/no questions and explicitly asks if the participant has someone who is making financial or healthcare decisions on their behalf (i.e., an active legal authorized representative). Once interest and eligibility are confirmed, participants are sent a PDF of the consent form to sign electronically using AdobeSign via Mayo Clinic’s Participant Tracking System (PTrax). PTrax is an institutional research software program designed to streamline the informed consent and re-consent processes, manage participant study status and history, track enrollments and accruals, populate the IRB Continuing Review form, and provide reporting and analytics as part of an institutional effort to transform the activation of clinical trials. Once participants provide their consent to partake in the study, they are sent a message in the patient portal with a link to the speech recording platform along with instructions. This study aims to use the data provided to predict neurological degenerative disease progression based on speech alone, as a means of developing an easy-to-use and cost-effective screening tool.

### Dataset

We exported longitudinal record data of participants invited to the speech capture study from March to July 2024 from Epic, Qualtrics, PTrax, and the recording platform. Socioeconomic status was assessed using the Housing-based Socioeconomic Status (HOUSES) index and the Area Deprivation Index (ADI) national rank. The HOUSES index is a practical and adaptable tool for assessing socioeconomic status using housing data. It effectively correlates with traditional socioeconomic status measures and predicts various health outcomes^17^. Higher HOUSES index values indicate higher socioeconomic status, while lower values indicate lower socioeconomic status. The ADI national rank measures neighborhood socioeconomic disadvantage across the United States, ranking neighborhoods based on various socioeconomic factors such as income, education, employment, and housing characteristics^18,19^. It is widely used in public health research to understand the impact of socioeconomic factors on health outcomes and access to healthcare services. ADI provides a score for each neighborhood, with higher scores indicating greater socioeconomic disadvantage.

The collected data included participants’ age, sex assigned at birth (hereafter referred to as sex), date of invitation to participate, recruitment process checkpoints along with their dates, devices used for speech recording on the platform, and any comments entered post-participation. Residence information was utilized to measure the HOUSES index and the national Area Deprivation Index (ADI), and to determine whether the patient resided in an urban area, rural area, or urban cluster. The study was approved by the Institutional Review Board (IRB).

### Analysis

The longitudinal time series data for each patient undergoing the participation process was standardized to align with specific checkpoints, as demonstrated in Figure 1. At each step, it was possible for patients to not respond to the research coordination team, which was defined as the “No Response” stage. After providing consent, participants were also free to withdraw it at any time. While most individuals followed one of the typical pathways depicted in Figure 1, there were 83 cases (1.4% of the total) in which participants deviated from these paths, necessitating intervention by research coordinators. These atypical cases were primarily attributed to personal circumstances or the involvement of a legally authorized representative.

**Figure 1.**
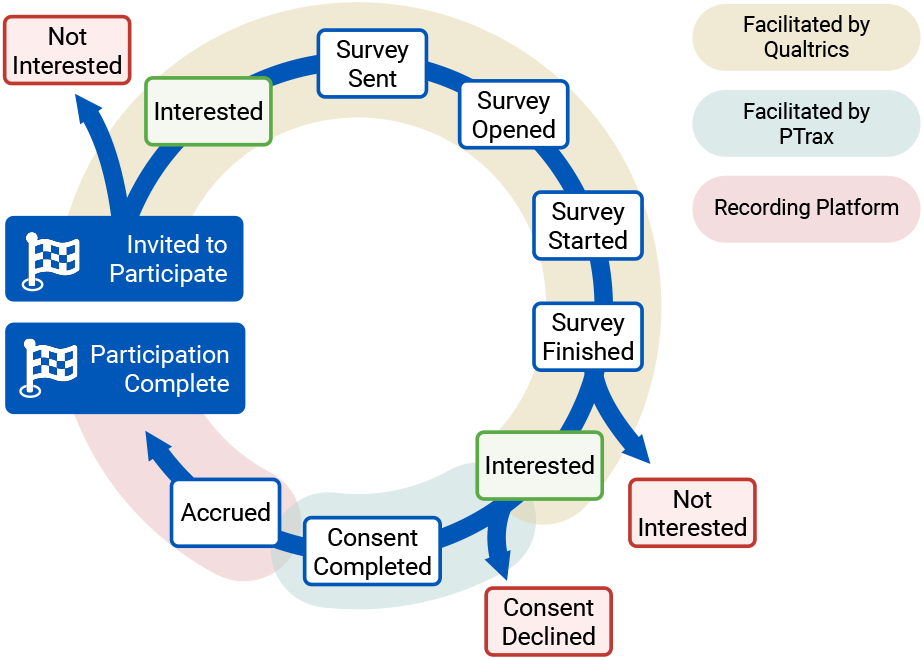
Simplified view of the participation enrollment

Kruskal-Wallis and Wilcoxon rank-sum tests were used to compare the median age, socioeconomic indices, and time taken to reach different steps of the study. Analysis was conducted at two levels: at each checkpoint and through an end-to-end investigation of participants who completed the study. For each path a participant could take at each checkpoint, age, ADI national rank, and HOUSES index were compared to identify statistically significant differences (defined as *p*<0.05). At the end-to-end level, additional comparisons were made across sex, population area (urban, rural, urban cluster), ADI national rank, and HOUSES index to evaluate differences in time taken to complete participation, from initial invitation to accrual, and whether participants completed the study, regardless of the path taken.

Since both Kruskal-Wallis and Wilcoxon rank-sum tests require at least five data points in each comparison category, pathways with fewer than five participants without missing values were excluded from the analysis. To further assess the nature of missing data across key sociodemographic variables, including age, ADI national rank, and HOUSES index, we conducted Little’s MCAR test^20^. This test evaluates whether data are missing completely at random (MCAR), which informs the appropriate handling strategy. Based on the results, we adopted a pairwise deletion approach for statistical analyses, allowing each test to include all available cases for the specific variable of interest. This method was chosen to preserve sample size and maintain statistical power, particularly in subgroups with limited data.

## Results

A total of 5,846 patients were invited to participate in the study between March and July 2024. Of these participants, 3,283 (56.2%) were female, 2,560 (43.8%) male, and 3 (0.05%) were unknown. Most participants (*n*=5,478, 93.7%) identified as White. The age distribution ranged from 18 to 96 years, with a median (IQR) age of 63 (48-72) years. Regarding geographic distribution, 56.5% of invited participants resided in urban areas, 23.3% in rural areas, and 20.2% in urban clusters (Table 1). Following accrual completion, participants utilized various devices to access the recording platform. Apple-based mobile devices (iPhone, iPad) were most frequently used (*n*=141, 34.0%), followed by Windows-based computers (*n*=134, 32.3%), Apple-based computers (*n*=82, 19.8%), Android-based mobile devices (*n*=52, 12.5%), and other devices (*n*=6, 1.4%).

**Table 1.** Demographics of the study participants.

| Category | Subcategory | Number of Participants (%) |
| --- | --- | --- |
| Sex | Male | 3283 (56.2%) |
|  | Female | 2560 (43.8%) |
|  | Unknown | 3 (0.0%) |
| Race | White | 5478 (93.7%) |
|  | Choose not to disclose | 65 (1.1%) |
|  | Black or African American | 53 (0.9%) |
|  | African American | 44 (0.8%) |
|  | Other | 206 (3.5%) |
| Age | 18 - 33 | 569 (9.7%) |
|  | 34 - 49 | 999 (17.1%) |
|  | 50 - 64 | 1607 (27.5%) |
|  | 65 - 80 | 2203 (37.7%) |
|  | 81 + | 468 (8.0%) |
| Population | Urban area | 3303 (56.5%) |
|  | Rural area | 1363 (23.3%) |
|  | Urban cluster | 1180 (20.2%) |
| Device used for participation | Apple-based mobile device | 141 (34.0%) |
|  | Windows-based computer | 134 (32.3%) |
|  | Apple-based computer | 82 (19.8%) |
|  | Android-based mobile device | 52 (12.5%) |
|  | Other devices | 6 (1.4%) |

The Area Deprivation Index (ADI) national rank of participants spanned the entire range from 1 to 100, with a median (IQR) of 44 (28-61), indicating representation across diverse socioeconomic backgrounds. Similarly, the Housing-based Socioeconomic Status (HOUSES) index percentile ranged from 1 to 100, with a median (IQR) of 70 (43-88), demonstrating considerable variability in housing conditions among participants. Neither age, ADI national rank, nor HOUSES index exhibited normal distribution according to the Anderson-Darling test (*p*<0.001 for all variables). Figure 2 illustrates the distribution of the HOUSES index percentile and ADI national rank.

**Figure 2.**
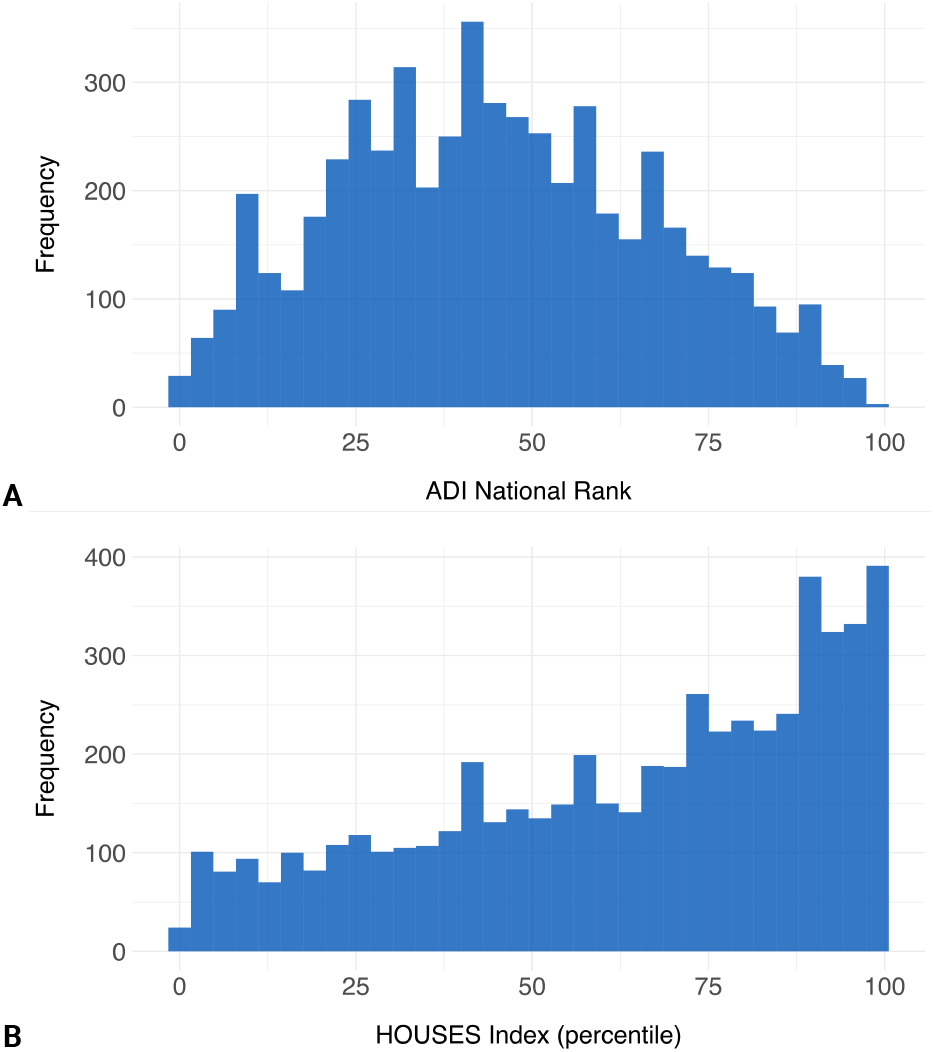
Histograms showing the distribution of (A) Area Deprivation Index (ADI) national rank and (B) Houses Index percentiles among study participants

To assess the nature of missing data, Little’s MCAR test was conducted on age, ADI national rank and HOUSES index and yielded a *p*-value of 0.243. The absence of statistical significance supported the assumption that the data were missing completely at random. This suggests that the missingness was likely not systematically related to other socioeconomic variables included in the analysis. Accordingly, pairwise deletion was applied in subsequent analyses to maximize data retention and ensure robust comparisons between study completers and non-completers.

### Comprehensive Participation Analysis

Significant differences were observed between participants who completed the study and those who did not across several demographic and socioeconomic factors. Age was higher among completers (median 66.4 years) compared to noncompleters (median 62.8 years; *p*<0.001), suggesting that older individuals were more likely to complete participation (Table 2). Participants who completed the study also resided in slightly less socioeconomically disadvantaged areas, as indicated by lower ADI national ranks (median 41.0 vs. 44.5; *p*=0.040), although no significant differences were found in HOUSES percentiles.

**Table 2.** Comparison of socioeconomic and demographic characteristics between study completers and non-completers. The Wilcoxon Rank-Sum test was applied to assess differences between groups.

| Variable | Group | *n* | Median (IQR) | *p*-value |
| --- | --- | --- | --- | --- |
| Age (years) | Participation Not Complete | 5431 | 62.9 (47.5 - 72.7) | <0.001 |
|  | Participation Complete | 415 | 66.4 (56.0 - 72.5) |  |
| ADI National Rank | Participation Not Complete | 5018 | 44.5 (28.0 - 62.0) | 0.040 |
|  | Participation Complete | 385 | 41.0 (27.0 - 56.0) |  |
| HOUSES Percentile | Participation Not Complete | 5052 | 70.0 (42.0 - 88.0) | 0.760 |
|  | Participation Complete | 387 | 71.0 (44.0 - 88.0) |  |

Sex differences in enrollment time were significant, with female participants taking longer to enroll than males (median 38.5 vs. 32.0 days; *p*=0.010) (Table 3). Additionally, geographic location influenced time to complete enrollment. Participants from urban areas enrolled more quickly than those from rural or urban cluster regions (median 32.0 vs. 41.0 and respectively; *p*=0.011). However, no significant differences were found in completion time across sex or geographic categories. No significant associations were detected between HOUSES indices and study completion. Additionally, no significant relationship was found between the device type used for task completion and the completion or participation time.

**Table 3.**
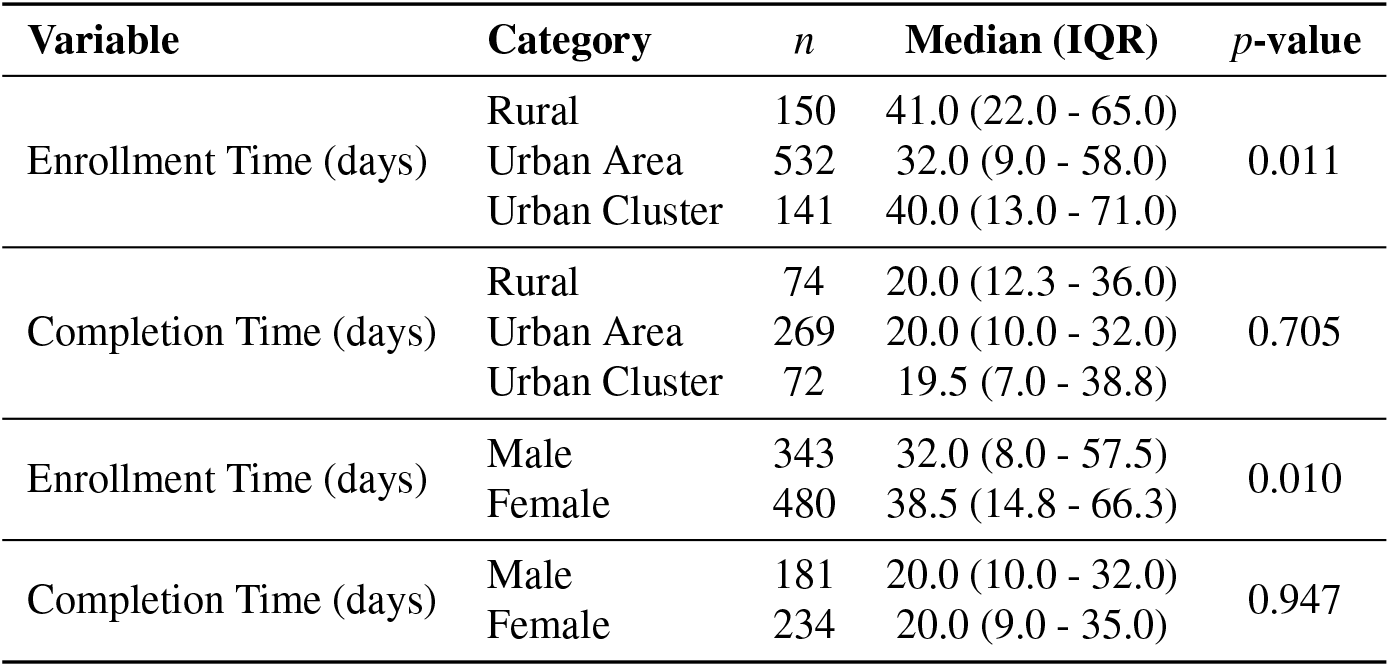
Comparison of enrollment and completion times stratified by population type and sex. Statistical significance was assessed using either the Wilcoxon Rank-Sum test or the Kruskal-Wallis test, depending on the number of categories.

| Variable | Category | *n* | Median (IQR) | *p*-value |
| --- | --- | --- | --- | --- |
| Enrollment Time (days) | Rural | 150 | 41.0 (22.0 - 65.0) | 0.011 |
|  | Urban Area | 532 | 32.0 (9.0 - 58.0) |  |
|  | Urban Cluster | 141 | 40.0 (13.0 - 71.0) |  |
| Completion Time (days) | Rural | 74 | 20.0 (12.3 - 36.0) | 0.705 |
|  | Urban Area | 269 | 20.0 (10.0 - 32.0) |  |
|  | Urban Cluster | 72 | 19.5 (7.0 - 38.8) |  |
| Enrollment Time (days) | Male | 343 | 32.0 (8.0 - 57.5) | 0.010 |
|  | Female | 480 | 38.5 (14.8 - 66.3) |  |
| Completion Time (days) | Male | 181 | 20.0 (10.0 - 32.0) | 0.947 |
|  | Female | 234 | 20.0 (9.0 - 35.0) |  |

### Step-by-Step Participation Analysis

The majority of invited participants either did not read/respond to the initial invitation via Epic (*n*=2,736) or expressed no interest (*n*=1,752). Among the 1,358 participants who initially expressed interest, 415 (30.6%) ultimately completed the study in its entirety. Throughout various stages of the recruitment process, a total of 3,346 participants failed to respond to follow-up communications from the research coordination team. Analysis of participant age across different pathways revealed that individuals who did not respond to the invitation or eligibility check were significantly younger than those who proceeded toward study completion. This age disparity contributed to an increase in the median age of participants completing the study (66.4 years) compared to the overall invited cohort (63.0 years). A similar pattern was observed among the 95 participants who withdrew consent after initially providing it but before being accrued for the recording session, with these participants having a median age of 55.8 years compared to 66.3 years for those who continued with the recording. While no significant differences in HOUSES index were observed across different participation pathways, participants who did not respond to the initial invitation had significantly higher ADI national ranks (median=45) compared to those who expressed interest (median=42), indicating residence in more socioeconomically disadvantaged neighborhoods. No other significant differences in ADI national ranks were observed across subsequent recruitment steps. Figure 3 provides a detailed breakdown of participants in each step and the paths they take.

**Figure 3.**
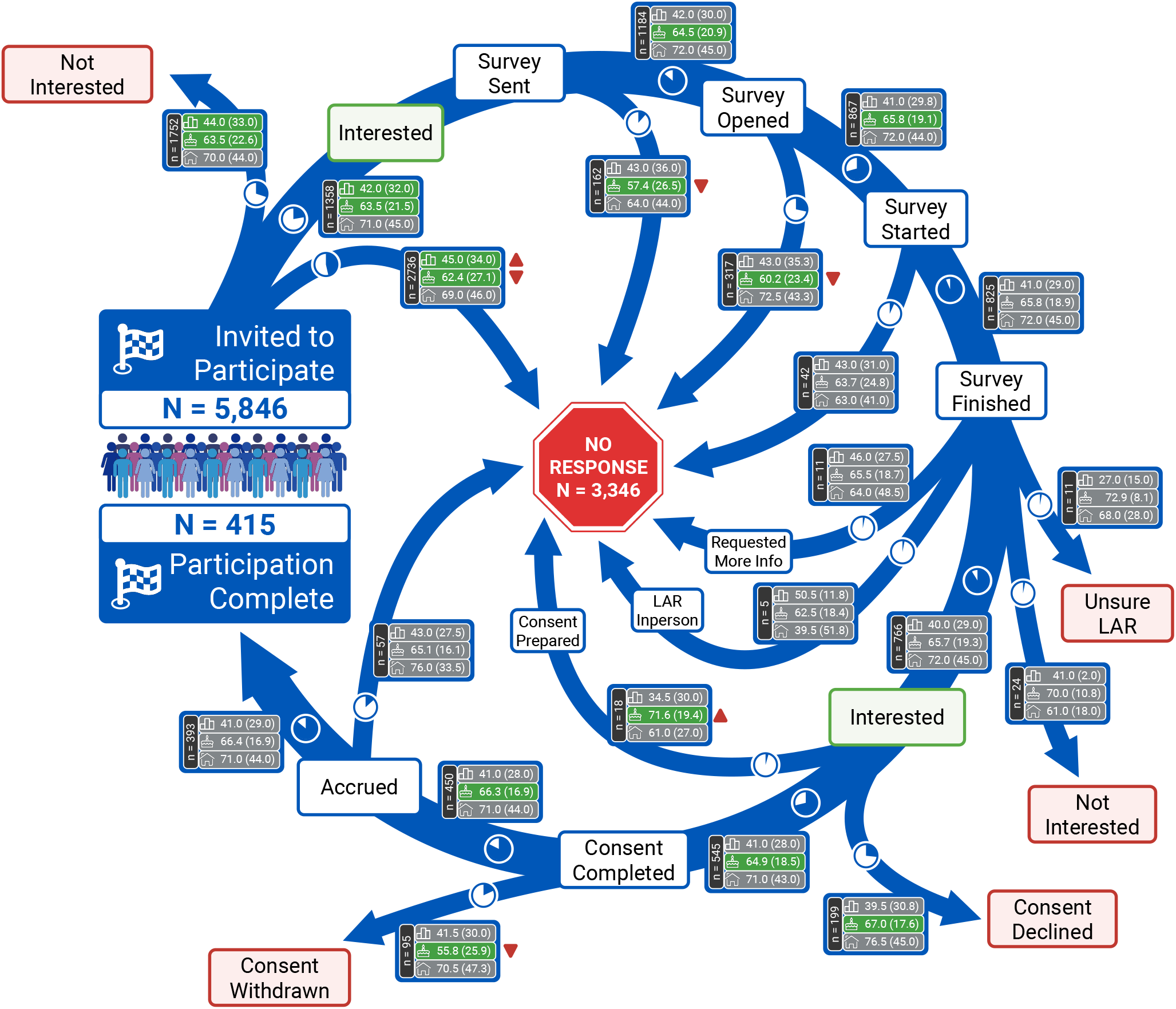
Step-by-step participation analysis. At each checkpoint in the participant journey, the median (IQR, interquartile range) of ADI, age, and HOUSES index are reported for participants following different paths. Significant differences (*p*<0.05) in the medians of these measures are highlighted with a green shade. Small pie charts on each path illustrate the relative frequency of participants at each stage. Paths with fewer than five participants are excluded from the analysis and are not displayed. LAR, Legally Authorized Representative.

## Discussion

This study provides a comprehensive analysis of participant recruitment pathways in a digital speech research study, revealing important associations between sociodemographic factors and participation outcomes. Understanding these pathways is critical for several reasons. First, recruitment is a fundamental challenge in clinical research, with participation rates in many medical studies remaining persistently low. Second, as digital recruitment methods become increasingly prevalent, it is essential to evaluate whether these approaches inadvertently perpetuate or even exacerbate existing disparities in research participation. Our findings demonstrate that systematic analysis of recruitment pathways can uncover patterns of participation and non-participation that may not be apparent when examining only final enrollment outcomes. Our results revealed several notable patterns in digital recruitment and engagement that challenge prevailing assumptions and highlights the need for targeted strategies to promote retention in remote search.

### Age-Related Participation Patterns

The significant age differences observed at various dropout points suggest that digital recruitment methods may be less effective for younger populations. This finding challenges the conventional wisdom that digital methods inherently appeal to younger participants and suggests that age-specific engagement strategies may be necessary throughout the recruitment process. While younger individuals may be more comfortable with technology, our results and others suggest that younger participants are more likely to disengage or drop out of digital studies over time^21,22^. This emphasizes the importance of age-sensitive retention strategies that extend beyond initial recruitment, including tailored messaging and incentives to support long-term engagement.

### Geographic Disparities in Enrollment Timing

The observation that participants from urban areas completed enrollment significantly faster than those from rural areas or urban clusters highlights potential geographic disparities in research accessibility. This finding aligns with broader concerns about the urban-rural divide in healthcare access and suggests that digital recruitment, while theoretically boundaryless, may still be influenced by geographic factors affecting infrastructure, digital literacy, or healthcare engagement^23,24^. This contrasts with some findings in telehealth adoption, which suggest that technology can overcome location-based care barriers^25^. Our results, however, indicate that digital research recruitment remains influenced by geographic factors, highlighting the need for strategies tailored to the rural digital context.

### Socioeconomic Disadvantage and Participation

Perhaps most notably, our analysis revealed that participants from neighborhoods with higher socioeconomic disadvantage (higher ADI national ranks) were significantly less likely to respond to initial invitations. This finding is consistent with prior research demonstrating that individuals from more disadvantaged areas experience greater barriers to engaging with digital health, including higher no-show rates and lower uptake of telehealth services^26,27^. This finding suggests that digital recruitment methods may perpetuate existing socioeconomic disparities in research participation if not specifically designed to address these barriers. The absence of significant differences in the HOUSES index across participation pathways, despite differences in the ADI national rank, suggests that neighborhood-level factors may play a more influential role in research participation than individual housing circumstances.

### Sex Differences in Enrollment Dynamics

Females took longer to complete the enrollment process than males, a difference that warrants further investigation. This may reflect differences in time availability, caregiving responsibilities, or engagement with digital health platforms that could impact recruitment strategies. While underexplored in the current literature, our finding encourages further investigation into potential influences of sex in digital research, which may inform tailored recruitment strategies and design of more flexible participation processes.

### Recruitment Funnel Attrition

Of 5,846 individuals invited, a large proportion did not respond or declined participation, and only 415 of all the invited individuals (7.1%) completed the study. This significant attrition is consistent with patterns observed in other digital recruitment efforts^28,29^. These findings highlight the need for iterative, multi-touch recruitment strategies that re-engage potential participants and address barriers to an active enrollment.

## Conclusions

This study demonstrates that digital recruitment methods in neurological research are subject to demographic, geographic and socioeconomic influences that affect the representativeness of study populations. Our findings suggest that while digital recruitment offers many advantages, including broader reach and efficiency, it does not automatically overcome traditional barriers to research participation and may introduce new ones. The digital divide appears to manifest in nuanced ways throughout the recruitment process, potentially influencing who participates in neurological research and, consequently, who benefits from its findings. The higher dropout rates among younger participants, those from more socioeconomically disadvantaged neighborhoods, and those in nonurban areas highlight the need for targeted strategies to engage these populations. The observed sex-based differences in enrollment speed also warrant further investigation, particularly regarding time burden or caregiving roles that may disproportionately impact female participants. The relatively low overall completion rate (7.1% of invited participants) underscores the persistent challenge of recruitment in specialized medical research, even with digital methods. Collectively, these findings reinforce the importance of refining digital recruitment strategies to bridge the persistent digital divide and promote research participation. Specific recommendations for addressing these challenges are summarized in Box 1.

### Box 1

**Recommendations for Enhancing Digital Recruitment in Medical Research**

Based on our findings, we propose the following strategies to improve inclusivity and effectiveness in digital recruitment:

- **Use Multi-Channel Approaches:** Combine digital methods with traditional outreach, especially for populations with limited online access (e.g., rural or socioeconomically disadvantaged communities).
- **Tailor Age-Specific Engagement:** Develop targeted messaging and user experiences for different age groups, with a focus on reducing dropout among younger participants.
- **Address Geographic Barriers:** Mitigate location-based disparities by offering technical support and alternative participation options for rural and urban clusters.
- **Track Recruitment Analytics:** Monitor recruitment data to identify dropout points and demographic trends, enabling real-time strategy adjustments.
- **Minimize Participation Burden:** Streamline enrollment to reduce time demands, benefiting participants with limited availability, particularly women.
- **Consider Socioeconomic Accessibility:** Ensure recruitment materials are inclusive and consider incentives or support to offset participation costs.
- **Optimize for Device Diversity:** While device type did not affect completion time, platforms should be mobile-friendly and accessible across devices.

Future research should explore the mechanisms behind demographic and socioeconomic disparities in recruitment and assess whether these patterns extend beyond neurological research. Intentional design and continuous evaluation are key to ensuring digital methods foster, rather than hinder, inclusivity in research.

## Data Availability

Due to the sensitive nature of patient data, the datasets are not publicly available but may be obtained from the corresponding author upon reasonable request and with appropriate institutional approvals.

## Acknowledgements

Some figures were created with BioRender.com

## Reprints and correspondence

Hugo Botha, 200 First St. SW Rochester, MN 55905

